# VULVOVAGINAL CANDIDIASIS: KNOWLEDGE, PRACTICES AND OCCURRENCE AMONG PREGNANT WOMEN RECEIVING ANTENATAL CARE IN A TEACHING HOSPITAL, GHANA

**DOI:** 10.1101/2024.12.12.24318968

**Authors:** Florence Shine Edziah, Princess Ruhama Acheampong, Philip Apraku Tawiah, Cedric Dzidzor Amengor, Godsway Edem Kpene, Grace Otobea Amponsah, Priscilla Appiah Baffoe, Georgina Korankye

## Abstract

**Background:** Vulvovaginal Candidiasis is a condition commonly caused by *Candida albicans*. It is the second most common infection of the female genitalia affecting many women worldwide. Studies have identified unhealthy genital care practices associated with the infection among women including expectant mothers. Knowledge of the various signs and symptoms is crucial for early detection, reporting, and treatment. Good knowledge may influence healthy practices limiting the infection and its complications. This study assessed the relationship between knowledge, practices and occurrence of Vulvovaginal candidiasis among pregnant women accessing antenatal care at a teaching hospital in Ghana.

**Methods:** A cross-sectional study was conducted among 336 pregnant women receiving antenatal care at the Ho Teaching Hospital. A structured questionnaire was employed in assessing their knowledge on the infection and some practices regarding vaginal hygiene. Hospital records of these participants were further checked to verify the occurrence of the infection among them. Analysis to identify associations between outcome variables and risk factors as well as significance level was carried out.

**Results:** Out of the 336 gestational mothers involved in the study, 27% were found to have been diagnosed with candidiasis at the time of the study. Pregnant women who usually use antibiotics had 2.25 increased odds of developing Vulvovaginal Candidiasis (VVC) compared to those who do not [OR:2.25 95CI:1.33-3.79; p-value = 0.003]. Again, a greater percentage of the study participants, 85% had good knowledge whiles 5% had poor knowledge.

**Conclusion:** The occurrence of VVC was elevated in the study jurisdiction. Frequent antibiotic use was found as a significant factor associated with the occurrence of the infection.

## INTRODUCTION

Vaginal Candidiasis has been reported globally as a condition afflicting a significant number of women. It is a global burden among women with 75% experiencing an episode in their lifetime (5). An investigation carried out on the prevalence of recurrent vulvovaginal candidiasis in five European countries and the U.S revealed that, over 40% of women reported at least four acute episodes of Vulvovaginal candidiasis in a 12-month period(1).

Studies show a rise in Vulvovaginal candidiasis in Asia and Africa (2). In South Asia, a prevalence of 55.5% was recorded among Pregnant women receiving care at a gynecological and obstetrics unit in a tertiary healthcare facility (3) whereas, in Africa, a recent study found a total prevalence of 43,.3% among pregnant women in South East Nigeria (Ekwealor et al., 2023)

In Ghana, specifically in the middle belt, a 36.5% prevalence was observed among pregnant women (4), this estimate is similar to a study conducted in the Ho Municipality (30.7%) (5). The disease is commonly caused by *Candida albicans* and it infests the vulva. Gestational mothers experience this condition due to the change in pH and sugar content of vaginal secretions resulting in the overgrowth of the causative organism (6). Apart from the biological causal pathway, lifestyle and hygiene practices have also been reported to influence the infection rate greatly (7).

Pregnant women indulge in some unhealthy practices as a result of inadequate knowledge, enhancing their vulnerability to candidiasis. A study by Nkamedjie Pete et al (2019) among antenatal care attendees on genital hygiene practices identified some risky activities such as the use of antiseptic solutions for genital cleansing and the use of synthetic underwear. Other findings have identified practices such as; not changing underwear daily, wearing dump underwear and using herbal combinations to wash the private part for easy baby delivery (8). Again, a survey on the analysis of genital hygiene behavior among women revealed that most pregnant women undergo vaginal douching (Karadeniz, Öztürk & Ertem, 2019).

Improper antibiotic use, an associated factor of increased prevalence has also been identified among gestational mothers (9). Among the risk factors, antibiotic use was ranked highest followed by intercourse, humid weather and use of feminine hygiene products (10). Antibiotics destroy the protective bacteria flora in the vagina and allow the overgrowth of yeast. Some findings indicate that 28-33% women who undergo antibiotic therapy experience symptomatic genital candidiasis (11). Short courses of oral antibiotic therapy have been identified with an increased prevalence of symptomatic Vulvovaginal Candidiasis (Shukla and Sobel, 2019). Several studies indicate a frequent occurrence of vulvovaginal candidiasis (VVC) following antibiotic use(12).

Knowledge of the various signs and symptoms is crucial for early detection, reporting, and treatment. As far as the knowledge of vaginal discharge is concerned, a common sign associated with the infection is abnormal discharge A study by Varghese et al (2017) found that, abnormal vaginal discharge was perceived by the majority of women as normal. This indicates a challenge in knowledge with regards to normal vaginal discharge among women in relation to the infection. A common gynecological complain among pregnant women is the inability to differentiate between normal and abnormal vaginal discharge leaving a gap in knowledge(13)

Knowledge on various signs such as abnormal vaginal discharge is key to early detection. Knowledge may influence healthy practices and enhance preventive measures hence limiting the rate of infection. Most studies on Vulvovaginal candidiasis worldwide, Africa and Ghana focused on prevalence. For instance, a prevalence of 30.7% was found in a study conducted at the Ho Teaching Hospital, Ghana. However, there was no further investigation to identify predisposing factors associated with such rate of occurrence. It is, therefore, relevant to assess the knowledge and practices of these women in relation to the rate of occurrence for appropriate intervention measures, hence this study seeks to assess the relationship between knowledge, practices and occurrence of VVC among the study participants at the Ho Teaching Hospital.

## Materials and Methods

### Study design, participants and setting

A cross-sectional study design was employed for this study.

This design was adopted to describe the distribution of candidiasis, knowledge levels, and practices among pregnant women across different demographic variables such as age, socioeconomic status, or education. It also helped in establishing the association between Knowledge, attitude and occurrence of the infection among the study group.

The study site is located in the Volta Region, Ho, Ghana and serves as the major referral center in the Region with 340-bed capacity. The facility provides specialized healthcare services such as Cardiothoracic services, intensive care, dialysis, dental services, operation smile, operation hernia, etc. Obstetrics and Gynecological services, pharmaceutical services, laboratory services, Ear, Nose and Throat services as well as Eyecare services are delivered by the facility. Obstetrics and Gynecological services are rendered each day at the facility. The unit has about twenty-seven midwives who provide out-patient and in-patient care on daily basis. Women in the municipality and beyond visit the facility to receive care. On the average, 20 pregnant women report to the Antenatal Care Unit on a daily.

### Sample size determination

The Cochran sample size estimation formula was used to obtain the appropriate sample size. The sample size was estimated based on a 95% confidence level with the prevalence of a recent study in the Ho Municipality (30.7%) (5). A total sample size of 345 was obtained with 5% non-response rate factored in.

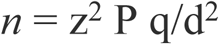

(*n* = Sample size, z = Standard normal deviation = 1.96 at 95% confidence limit, P = Prevalence rate = 31%, q = 1-P = 1–0.31% =0.69, d = Error margin = 5%)

### Sampling Procedure

A simple random sampling method was used to recruit pregnant women receiving antenatal care at the facility. All participants were gestational mothers who agreed to partake in the study. Each received a pre-enrollment briefing about the study, an informed written consent was obtained. Participants were guided to complete a pre-tested questionnaire to provide information nonfactors such as: age, marital status, occupation, educational level, knowledge on candidiasis and the presence or absence of genital symptoms.

### Inclusion and Exclusion Criteria

#### Inclusion Criteria

Pregnant women who provided informed consent after obtaining information on the study’s objectives, risks, and benefits.

Participants confirmed pregnant, either through formal records, a healthcare provider or self-reported.

Participants receiving antenatal care services at the period of the study.

Participants ability to communicate effectively in the language used for the study to ensure they can understand and respond to questions about candidiasis knowledge and practices accurately.

#### Exclusion Criteria

Pregnant women who did not provide informed consent.

Gestational mothers who do not speak or understand the language used in the study (and for whom translation services were not available) were excluded to avoid misunderstandings or misinterpretations of the questions and responses.

#### Data collection

Pregnant women attending ANC were approached, informed about the study, and asked to provide informed consent if willing to partake in the study. After the consent, a structured close-ended questionnaire was used to gather data on the sociodemographic characteristics (age, sex, marital status, level of education and occupation) of study participants. The questionnaire was also used to gather information on various aspects of knowledge and practices related to candidiasis. Questions covered symptoms, risk factors, prevention strategies and treatment options. Participants completed questionnaire in a private setting to ensure confidentiality. Again, the hospital records of these participants were further checked to verify the occurrence of the infection among them. Data on previous VVC diagnoses and related treatments was extracted from the facility’s records. Standardized data collection procedures were followed. The study was approved by the relevant ethical review board before the commencement of data collection. The data collection began on 5^th^ September 2023 after approval had been obtained from the facility to commence the study. The data collection ended on 12^th^ October 2023.

#### Data management and analysis

The administered questionnaires were appropriately coded and entered into Microsoft Excel 2013 for cleaning. Upon validation, the cleaned data was exported to STATA (statistical analysis software) Version16 for statistical analysis.

A “Yes” or “No” response provided to questions on knowledge was coded. A “Yes” response was scored “1” while a “No” response scored “0”. A sum of these scores represented the total score of respondents on the knowledge of Vulvovaginal candidiasis. Respondents who scored below 5 were classified as having poor knowledge. Scores above 5 but below 8 was scored as having moderate knowledge and scores above 8 was classified as good.

The various practices were recorded. Prevalence of vulvovaginal candidiasis was determined based on respondents’ expression of signs and symptoms associated with vulvovaginal candidiasis coupled with evidence from their medical records on laboratory investigations to establish the presence of the infection.

Categorical variables were presented as frequencies and percentages, while continuous variables were represented as means, median, standard deviations or interquartile ranges. “Bivariate analysis was performed to examine the correlation between Knowledge, practices and the outcome of interest at a significance level of p<0.05.

## RESULTS

### 3.0 Socio-demographic characteristics of gestational mothers

The table 1 below summarizes the socio-demographic characteristics of gestational mothers receiving antenatal care at the Ho Teaching Hospital at the time of the study. Out of the 336 participants, 196(58.33%) aged 30-39 years. In relation to education and employment,156(46.4%) each out of the total respondents had tertiary education and were self-employed. The greatest number of expectant mothers, 197(58.63%) were in the third trimester of their pregnancy. Majority of these mothers were Christians 317(94.4%) and married 263(78.3%).

**Table 1:**
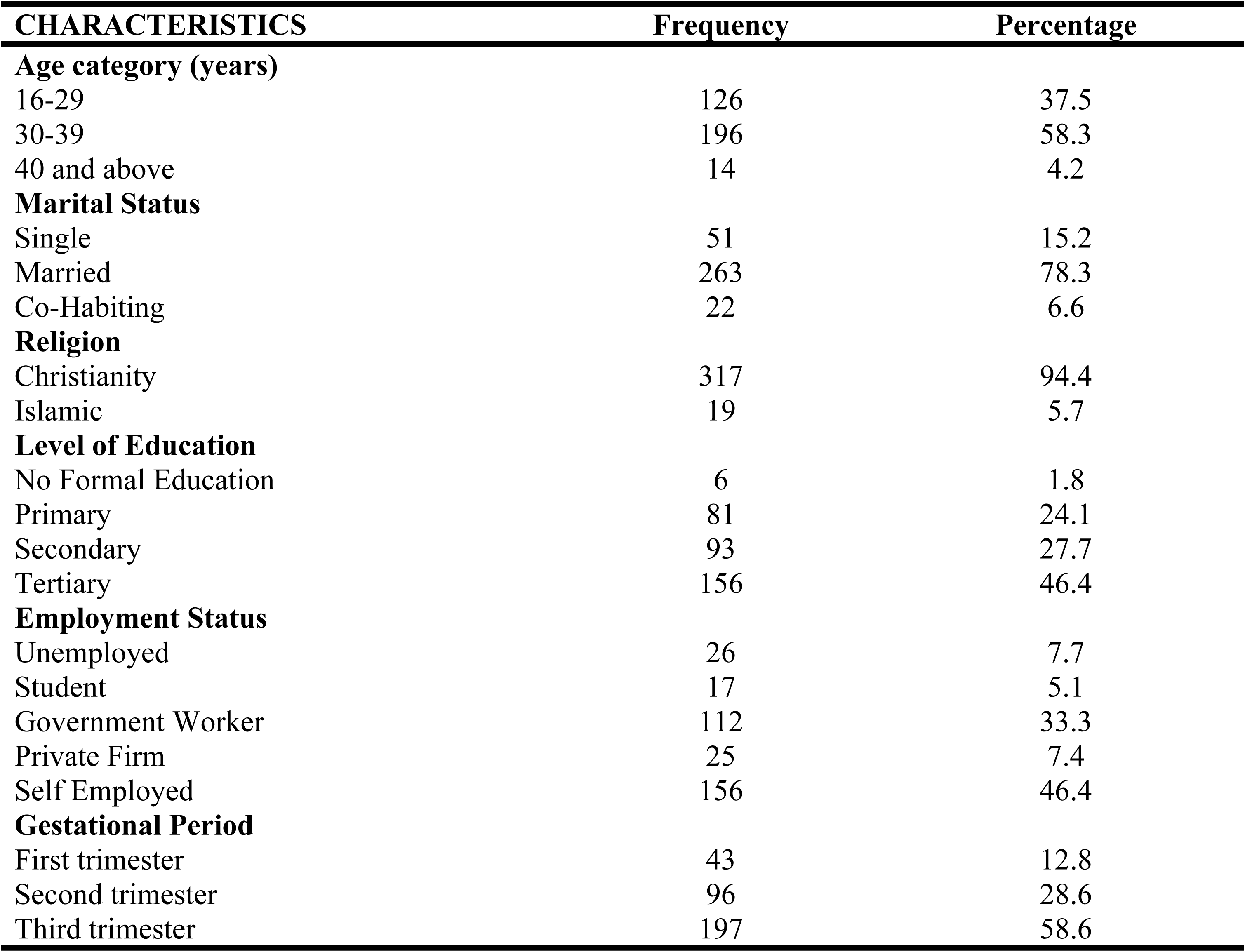
Socio-demographic characteristics of gestational mothers.

### 3.1 Knowledge of Pregnant Women on Vulvovaginal Candidiasis

Table 2 and Fig 1 below summarizes the level of knowledge among the study participants on the infection. A good knowledge was recorded among 80% on major signs and symptoms, however, it was described as a bacterial infection by 209(62.2%). Vomiting and fatigue was rightly indicated by 298(88.7%) of the respondents as not being a sign/symptom associated with the infection. burning sensation and pain in the vagina were rightly indicated as symptoms by 255(75.89%) and 242(72.02%) respectively. Vaginal itching was indicated by 308(91.67%) as a symptom of candidiasis. A cheese-like vaginal discharge with odor was indicated by majority, 300(89.29%) as the abnormal discharge associated with the infection. Almost all the respondents, 308(91.67%) viewed Vulvovaginal candidiasis as a possible infection that can occur in pregnancy. Generally, the knowledge level of the expectant mothers was good. A greater percentage of the respondents, 85% had good knowledge whiles 5% had poor knowledge.

**Table 2:** Knowledge of Pregnant Women on Vulvovaginal Candidiasis.

| Characteristics | Frequency | Percentage |
| --- | --- | --- |
| **Type of Infection** |  |  |
| Fungal | 76 | 22.6 |
| Bacterial | 209 | 62.2 |
| Viral | 21 | 6.3 |
| Parasitic | 30 | 8.9 |
| **Signs/Symptoms** |  |  |
| Vomiting |  |  |
| No | 298 | 88.7 |
| Yes | 38 | 11.3 |
| **Fatigue** |  |  |
| No | 283 | 84.2 |
| Yes | 53 | 15.8 |
| **Burning Sensation** |  |  |
| No | 81 | 24.1 |
| Yes | 255 | 75.9 |
| **Pain in Vagina** |  |  |
| No | 94 | 28.0 |
| Yes | 242 | 72.0 |
| **Vagina Itching** |  |  |
| No | 28 | 8.3 |
| Yes | 308 | 91.7 |
| **Normal Vaginal discharge** |  |  |
| No | 269 | 80.1 |
| Yes | 67 | 19.9 |
| **Abnormal discharge** |  |  |
| No | 43 | 12.8 |
| Yes | 293 | 87.2 |
| **Pain/discomfort during sex** |  |  |
| No | 106 | 31.6 |
| Yes | 230 | 68.5 |
| **Associated Vaginal discharge** |  |  |
| Normal without odor | 36 | 10.7 |
| Cheese-like with odor | 300 | 89.3 |
| **View on VVC in pregnancy** |  |  |
| **VVC cannot occur in pregnancy** |  |  |
| False | 308 | 91.7 |
| True | 28 | 8.3 |

*[Figure 1. Knowledge level of participants on VVC]*

**Figure 1.**
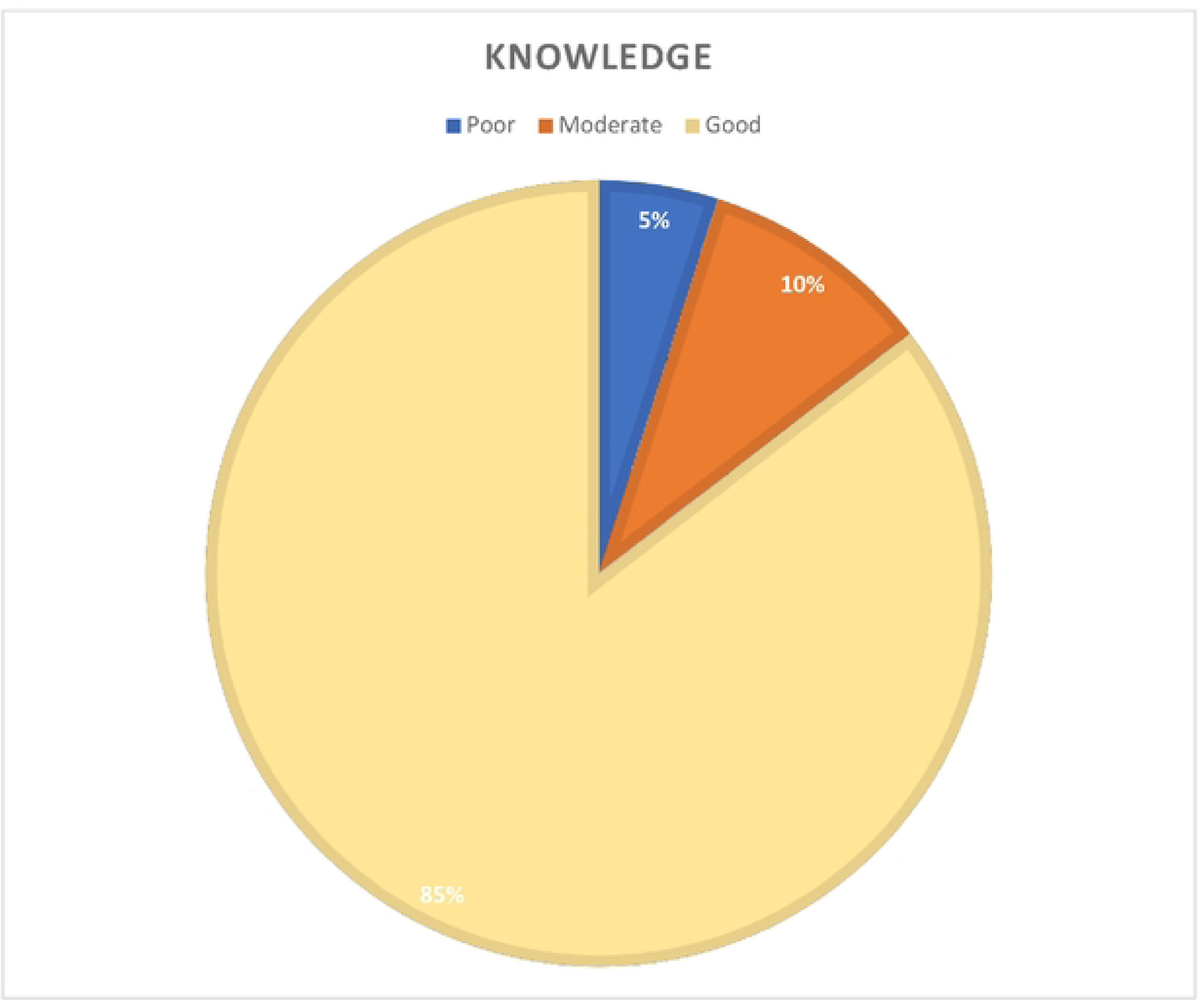
Knowledge level of participants on VVC.

### 3.2 Practices of Pregnant Women towards the occurrence of VVC

In Table 3 below, the summary of some practices among gestational mothers of this study can be found. The use of feminine hygiene washes 228(85.71%) and daily perineal pads223(66.37) was identified among the expectant mothers. Out of the total number of respondents, 302(89.88%), 321(95.54%), 244(72.62%) and 208(64.20%) changed their underwear daily, preferred cotton as fabric for their underwear, dried panties under the sun after washing and ironed their underwear respectively. Frequent antibiotic usage was indicated by 134(39.88%) of the study participants.

**Table 3:**
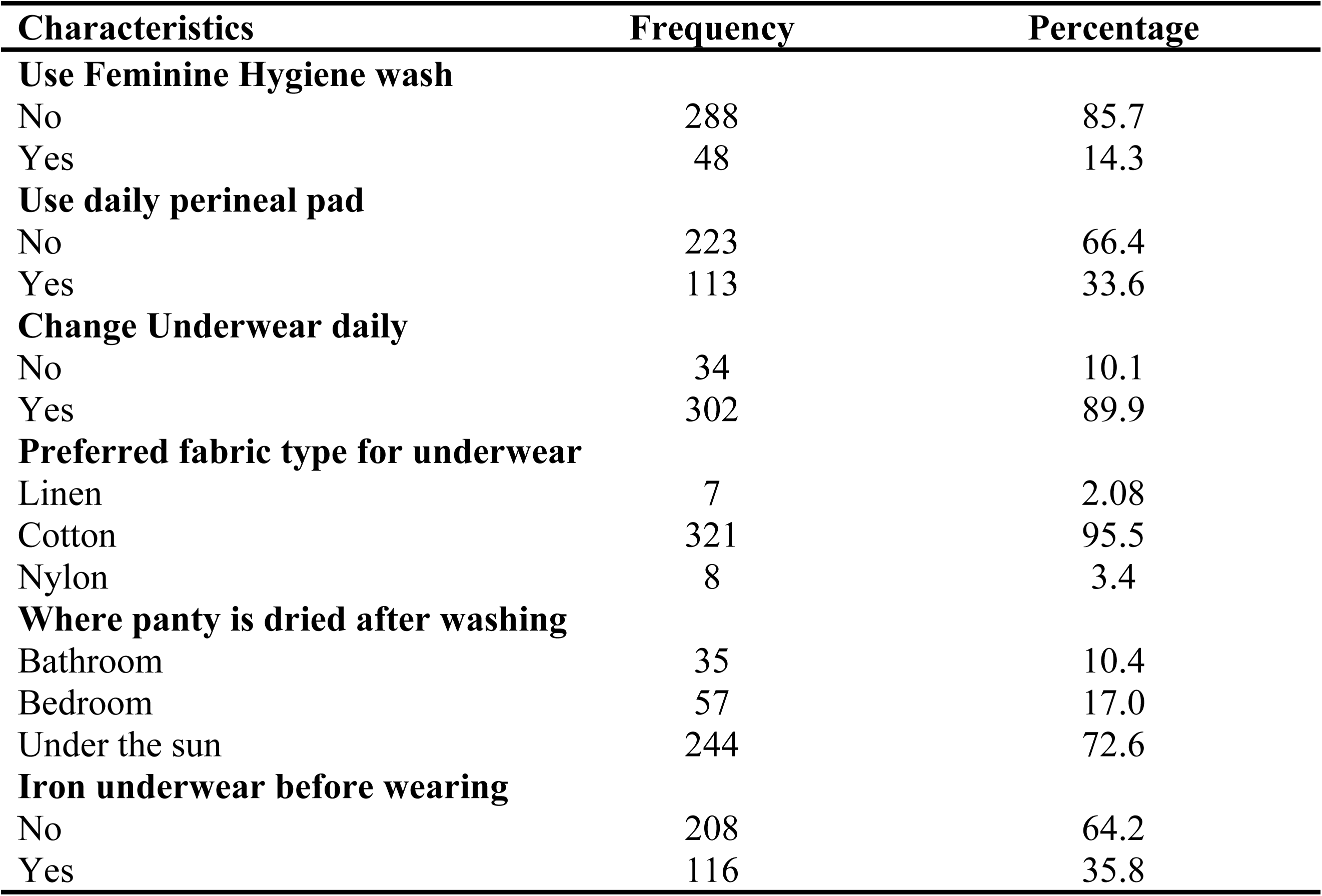
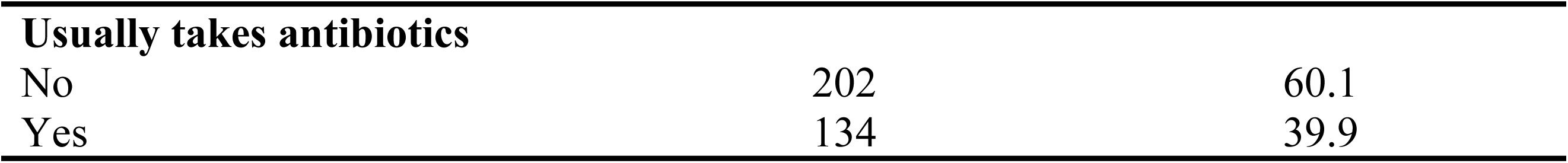
Practices of Pregnant Women towards the occurrence of VVC.

### 3.3 Occurrence of VVC among the Gestational Mothers

A prevalence of 27% was observed among the expectant mothers receiving antenatal care at the Teaching Hospital as shown in Fig 2 below.

**Figure 2:**
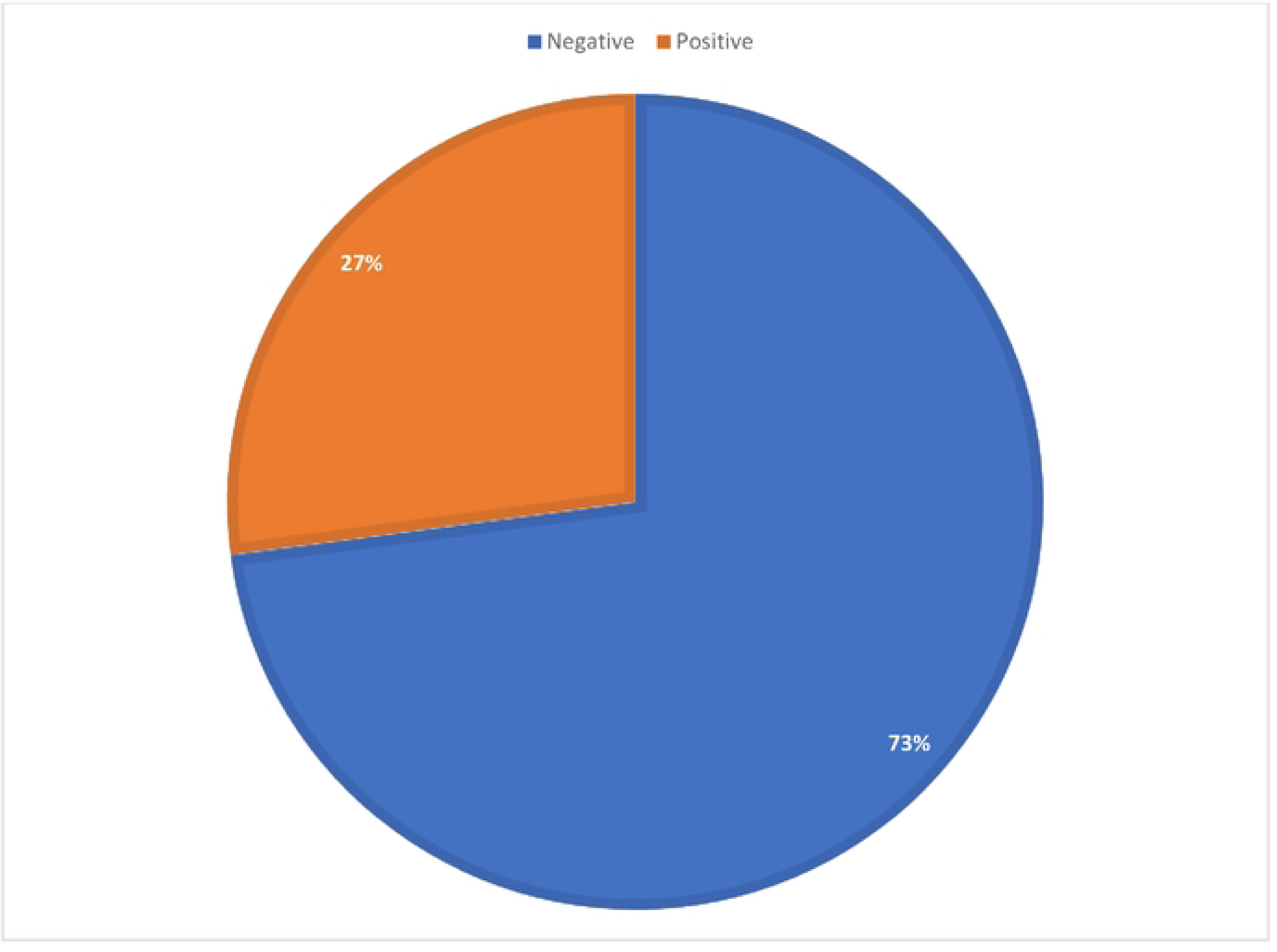
Prevalence of Vulvovaginal Candidiasis.

### 3.4 Effect of Socio-demographic characteristics on the occurrence of VVC

The table below describes the association between the various socio-demographic characteristics and the prevalence of the infection.

**Table 3:** Association between occurrence and socio-demographic variables.

| Characteristics | Total (%) | Confirmed cases |  | X<sup>2</sup> | p-value |
| --- | --- | --- | --- | --- | --- |
|  |  | Negative | Positive |  |  |
| **Age** |  |  |  |  |  |
| 16-29 | 126(37.5) | 84(66.67) | 42(33.33) | 4.02 | 0.134 |
| 30-39 | 196(58.33) | 150(76.53) | 46(23.47) |  |  |
| 40 and above | 14(4.17) | 11(78.57) | 3(21.43) |  |  |
| **Marital Status** |  |  |  |  |  |
| Single | 51(15.18) | 37(72.55) | 14(27.45) | 0.28 | 0.868 |
| Married | 263(78.27) | 193(73.38) | 70(26.62) |  |  |
| Co-habiting | 22(6.55) | 15(68.18) | 7(31.82) |  |  |
| **Religion** |  |  |  |  |  |
| Christianity | 317(94.35) | 232(73.19) | 85(26.81) | 0.21 | 0.650 |
| Islamic | 19(5.65) | 13(68.42) | 6(31.58) |  |  |
| **Highest level of education** |  |  |  |  |  |
|  | 6(1.79) | 4(66.67) | 2(33.33) | 3.78 | 0.287 |
| No Formal Education | 81(24.11) | 61(75.31) | 20(24.69) |  |  |
| Primary | 93(27.68) | 61(65.59) | 32(34.41) |  |  |
| Secondary | 156(46.43) | 119(76.28) | 37(23.72) |  |  |
| Tertiary |  |  |  |  |  |
| **Employment Status** |  |  |  |  |  |
|  | 26(7.74) | 17(65.38) | 9(34.62) | 2.49 | 0.647 |
| Unemployed | 17(5.06) | 12(70.59) | 5(29.41) |  |  |
| Student | 112(33.33) | 87(77.68) | 25(22.32) |  |  |
| Government worker | 25(7.44) | 17(68.00) | 8(32.00) |  |  |
| Private firm worker | 156(46.43) | 112(71.79) | 44(28.21) |  |  |
| Self-employed |  |  |  |  |  |
| **Gestational Period** |  |  |  |  |  |
|  | 43(12.8) | 29(67.44) | 14(32.56) | 0.99 | 0.608 |
| First trimester | 96(28.57) | 69(71.88) | 27(28.13) |  |  |
| Second trimester | 197(58.63) | 147(74.62) | 50(25.38) |  |  |
| Third trimester |  |  |  |  |  |

### 3.5 Effect of Knowledge and practices on the occurrence of VVC

A significant association was found between antibiotic use and the occurrence of vulvovaginal candidiasis as shown in Tab. 4 below, [***χ*2** = 7.2077, p-value = 0.007]

**Table 4.** Association between knowledge, practices and occurrence.

| Characteristics | Confirmed cases | | | $\chi^2$ | p-value |
| --- | --- | --- | --- | --- | --- |
|  | Total (%) | Negative | Positive |  |  |
| **Knowledge** |  |  |  |  |  |
| Poor | 27(8.04) | 19(70.37) | 8(29.63) | 0.98 | 0.613 |
| Moderate | 53(15.77) | 36(67.92) | 17(32.08) |  |  |
| Good | 256(76.19) | 190(74.22) | 66(25.78) |  |  |
| **Practices** |  |  |  |  |  |
| **Use feminine wash** |  |  |  |  |  |
| No | 288(85.71) | 210(72.92) | 78(27.08) | 0.00 | 1.000 |
| Yes | 48(14.29) | 35(72.92) | 13(27.08) |  |  |
| **Use perineal pads** |  |  |  |  |  |
| No | 223(66.37) | 165(73.99) | 58(26.01) | 0.39 | 0.534 |
| Yes | 113(33.63) | 80(70.80) | 33(29.20) |  |  |
| **Change underwear daily** |  |  |  |  |  |
| No | 34(10.12) | 23(67.65) | 11(32.35) | 0.53 | 0.466 |
| Yes | 302(89.88) | 222(73.51) | 80(26.49) |  |  |
| **Underwear fabric type** |  |  |  |  |  |
| Linen | 7(2.08) | 5(71.43) | 2(28.57) | 0.03 | 0.987 |
| Cotton | 321(95.54) | 234(72.90) | 87(27.10) |  |  |
| Nylon | 8(3.38) | 6(75.92) | 2(25.00) |  |  |
| **Drying of underwear** |  |  |  |  |  |
| Bathroom | 35(10.420) | 24(68.57) | 11(31.43) | 5.52 | 0.063 |
| Bedroom | 57(16.96) | 35(61.40) | 22(38.60) |  |  |
| Under the sun | 224(72.62) | 186(76.23) | 58(23.77) |  |  |
| **Iron Panty before use** |  |  |  |  |  |
| No | 208(64.20) | 157(75.48) | 51(24.52) | 2.05 | 0.152 |
| Yes | 116(35.80) | 79(68.10) | 37(31.90) |  |  |
| **Usually take antibiotics** |  |  |  |  |  |
| No | 202(60.12) | 158(78.22) | 44(21.78) | 7.21 | 0.007 |
| Yes | 134(39.88) | 87(64.93) | 47(35.07) |  |  |

### 3.6 Risk Factors associated with the occurrence of VVC

Table 5 provides a summary of a bivariate and multiple logistic regression of risk factors and Occurrence of VVC. Frequent antibiotic use was found to be a significant factor in the occurrence of VVC. Gestational mothers who usually use antibiotics had 2.25 increased odds of developing Vulvovaginal Candidiasis (VVC) compared to those who do not [OR:2.25 95CI:1.33-3.79; p-value = 0.003].

**Table 5.** Bivariate and multiple logistic regression of risk factors and Occurrence of VVC.

| Characteristics | OCCURRENCE OF VVC (n=336) |  |  |  |  |
| --- | --- | --- | --- | --- | --- |
|  | No. (%) | COR (95% CI) | p-value | AOR (95% CI) | p-value |
| **Age** |  |  |  |  |  |
| 16-29 | 126(37.5) | 1 |  | 1 |  |
| 30-39 | 196(58.33) | 0.61 (0.31-1.01) | 0.054 | 0.64(0.37-1.11) | 0.114 |
| 40 and above | 14(4.17) | 0.55 (0.14-2.06) | 0.371 | 0.64(0.16-2.59) | 0.536 |
| **Gestational Period** |  |  |  |  |  |
| First Trimester | 43(12.8) | 1 |  | 1 |  |
| Second Trimester | 96(28.57) | 0.81(0.37-1.76) | 0.597 | 0.91(0.41-2.04) | 0.819 |
| Third Trimester | 197(58.63) | 0.70(0.34-1.43) | 0.336 | 0.82(0.39-1.72) | 0.597 |
| **Employment status** |  |  |  |  |  |
| Unemployed | 26(7.74) | 1 |  | 1 |  |
| Student | 17(5.06) | 0.79(0.21-2.94) | 0.722 | 0.86(0.22-3.34) | 0.818 |
| Government worker | 112(33.33) | 0.54(0.22-1.37) | 0.194 | 0.60(0.22-1.66) | 0.346 |
| Private firm | 25(7.44) | 0.89(0.28-2.80) | 0.843 | 0.99(0.29-3.34) | 0.930 |
| Self- employed | 156(46.43) | 0.74(0.31-1.79) | 0.506 | 0.89(0.35-2.25) | 0.861 |
| **Frequent antibiotics use** |  |  |  |  |  |
| No | 202(60.12) | 1 |  | 1 |  |
| Yes | 134(39.88) | 1.94(1.19-3.16) | 0.008 | 2.25(1.33-3.79) | 0.003 |
| **Knowledge on VVC** |  |  |  |  |  |
| Poor | 27(8.04) | 1 |  | 1 |  |
| Moderate | 53(15.77) | 1.12(0.41-3.07) | 0.823 | 0.84(0.29-2.40) | 0.837 |
| Good | 256(76.19) | 0.83(0.34-1.97) | 0.666 | 0.75(0.30-1.85) | 0.593 |

\* Significant variable (p-value < 0.05)

## DISCUSSION

The study examined the knowledge, practices and prevalence of candidiasis among expectant mothers. It identified a relationship between lifestyle and hygienic practices with the occurrence of vaginal thrust. Some practices such as the use of feminine hygiene wash, perineal pads, the use of antibiotics among others. There was a strong association between frequent antibiotic use and the occurrence of candidiasis. The prevalence of candidiasis was identified from the expressions of participants and confirmations from their medical records. A 27% prevalence was observed with a link to antibiotic use, however, there was no relationship found between knowledge and some other practices in relation to the infection.

There was a slightly elevated 27% prevalence recorded compared to the 25% reported at Yemen(14), However, lower than the 29.2% prevalence indicated in a systematic review and meta-analysis conducted in Africa (Osman Mohamed et al., 2022) and the 30.7% recorded in the Ho Municipality among gestational mothers (5). The dichotomy in these observed prevalences could be associated with the difference in sample size, study design and geographical location.While this study engaged 336 participants, that which was carried out in Yemen and Ho Municipality involved 250 and 176 study subjects respectively. Given that VVC is common illness among pregnant women, there is a need to heighten awareness and screening for such mothers in the current study jurisdiction. This will foster early case detection and treatment to aid the mitigation of associated discomfort and the risk of complication during pregnancy.

An association was found between antibiotic use and the occurrence of vulvovaginal candidiasis. This conforms with some recent studies which found associations between antibiotic usage and the prevalence of the infection. A study by Dou (2014), indicated that the widespread use of antibiotics contributes to an increased prevalence of Candida vaginal infection. Their study identified pregnancy, diabetes mellitus, contraceptive and antibiotic use as predisposing factors for the infection (15). Also, reports from another study indicated that about 28-33% of women put on antibiotic therapy develop symptomatic genital candidiasis (11). Again, a retrospective study conducted Between 2014 and 2016, revealed that the use of gynecological antibiotics, systemic antibiotics, oral contraceptives as well as vaginal contraceptives were associated with an increase in the risk of vulvovaginal candidiasis (1). Another study characterizes antibacterial therapy, whether administered systemically or locally to the vagina, as the most common and predictable cause or triggering factor for symptomatic vulvovaginal candidiasis (VVC). (16). It is therefore evident that, antibiotic use has an impact on the occurrence of vulvovaginal candidiasis in most cases Hence, patients must be educated on the potential side effects of antibiotics, including the risk of candidiasis, and encourage them to take antibiotics only as prescribed and to complete the full course. Again, surveillance of antibiotic-resistant bacteria and their impact on infections such as candidiasis must be Strengthened. Monitoring resistance trends helps tailor guidelines for antibiotic use more effectively.

The study did not find the use of perineal pads, feminine hygiene wash, regular changing of underwear, use of cotton underwear, drying underwear under the sun and ironing of underwear to be significantly associated with Vulvovaginal candidiasis infection among the study participants. “This result aligns with the results of a study on the impact of supportive nursing instructions on the recurrence of vulvovaginal candidiasis infection during pregnancy. Their research similarly showed no notable association between practices such as using daily protective pads, wearing cotton underwear, changing wet underwear frequently, employing vaginal douching, performing vaginal douching during menses, and the recurrence of vulvovaginal candidiasis infection during pregnancy.(17). This could be due to the fact that Pregnancy significantly alters hormone levels, particularly estrogen and progesterone. These hormonal changes can make the vaginal environment more conducive to the overgrowth of Candida species, which is the primary cause of VVC. Such biological factors can outweigh the influence of personal hygiene practices, making them less significant in determining recurrence.

The question on the causative organism, thus fungi was missed by majority, 62.20% of the study participants referred to it as a bacterial infection though a good knowledge was recorded with other questions on knowledge. Burning sensation, Itchy vagina and abnormal vaginal discharge was rightly associated by the study participants with the infection. Questions on signs and symptoms were answered correctly. On the contrary, a study recorded poor knowledge on the signs and symptoms among pregnant women (18).

Findings on knowledge, practices, and prevalence of candidiasis among pregnant women emphasize the necessity of addressing this widespread but often ignored condition in maternal healthcare. Our findings revealed that though there is good knowledge on the signs and symptoms there still remains a gap on the knowledge of the causative agent.

## LIMITAIONS OF THE STUDY

It is impossible to draw causal relations between factors and the occurrence of VVC due to the fact that the study was cross sectional. Furthermore, questions on practices were self-reported which may be prone to recall bias.

## CONCLUSION

A high prevalence of VVC (27%) was reported among the expectant mothers visiting Ho Teaching Hospital for Antenatal Care. The findings thus suggests that candidiasis is a common illness among pregnant women, necessitating heightened vigilance in prenatal care and the creation of evidence-based therapeutic guidelines. Generally, these mothers had good knowledge on VVC, although most could not associate VVC as a fungal infection. Finally, only the frequent use of antibiotics was identified as a significant factor to VVC. Given that the frequent use of antibiotics is associated with VVC, expectant mothers should be educated on the dangers of the frequent use of antibiotics. Additionally, when administering antibiotics, healthcare providers choose narrow-spectrum antibiotics to reduce their impact on the entire vaginal microbiome. Further qualitative study should be undertaken to unearth the plausible drivers to expectant mothers in frequent utility of antibiotics. There is therefore the need for improved policies on healthcare practices and training programs aimed at pregnant women.

## Data Availability

Data will be made available upon reasonable request.

## Ethical consideration

The research protocol was approved by the Committee on Human Research Publication and Ethics (CHRPE) at Kwame Nkrumah University of Science and Technology, Kumasi, with the reference number CHRPE/AP /703/2 and permission was obtained from the Research, Policy, Planning, Monitoring, and Evaluation (RPPME) of the Ho Teaching Hospital Prior to the commencement of the study. A comprehensive information was provided to study participants concerning potential risks and benefits, privacy, confidentiality, data storage and usage, voluntary consent and withdrawal, compensation, declaration of conflicts of interest, and research funding. Subsequently, participants were required to carefully review and complete a written informed consent form before participating in the study.

## DECLARATION

All authors declare no conflict of interest.

## REFERENCES

1. Jacob L, John M, Kalder M, Kostev K. Prevalence of vulvovaginal candidiasis in gynecological practices in Germany: A retrospective study of 954,186 patients. Current medical mycology. 2018;4(1):6–11.

2. Disha T, Haque F. Prevalence and Risk Factors of Vulvovaginal Candidosis during Pregnancy: A Review. Infectious diseases in obstetrics and gynecology. 2022;2022:6195712.

3. Yadav LK, Yadav R. Prevalence of vaginal yeast infections in pregnant and non-pregnant women attending at Gynecology and Obstetrics Department of the tertiary care center in Central region of Nepal. Microbes and Infectious Diseases. 2023;4(1):225–30.

4. Konadu DG, Owusu-Ofori A, Yidana Z, Boadu F, Iddrisu LF, Adu-Gyasi D, et al. Prevalence of vulvovaginal candidiasis, bacterial vaginosis and trichomoniasis in pregnant women attending antenatal clinic in the middle belt of Ghana. BMC Pregnancy Childbirth. 2019;19(1):341.

5. Waikhom SD, Afeke I, Kwawu GS, Mbroh HK, Osei GY, Louis B, et al. Prevalence of vulvovaginal candidiasis among pregnant women in the Ho municipality, Ghana: species identification and antifungal susceptibility of Candida isolates. BMC Pregnancy and Childbirth. 2020;20(1):266.

6. Tan BH, Chakrabarti A, Li RY, Patel AK, Watcharananan SP, Liu Z, et al. Incidence and species distribution of candidaemia in Asia: a laboratory-based surveillance study. Clinical microbiology and infection : the official publication of the European Society of Clinical Microbiology and Infectious Diseases. 2015;21(10):946–53.

7. Ghaddar N, El Roz A. Emergence of Vulvovaginal Candidiasis among Lebanese Pregnant Women: Prevalence, Risk Factors, and Species Distribution. 2019;2019:5016810.

8. Nun D, Adesuyi E, Olawoore S. Knowledge Attitude and Practices of Pregnant Women Attending Comprehensive Health Centre, Isolo, Ondo State towards Hygienic Practice. International Journal of Tropical Disease & Health. 2018;30:1–10.

9. Christopher MA, Nyoyoko VF, Antia UE, Eyo IE. Prevalence of vulvovaginal candidiasis in pregnant women attending antenatal clinic in Abak, South-South Nigeria. Microbes & Infectious Diseases. 2022;3(3).

10. Yano J, Sobel JD, Nyirjesy P, Sobel R, Williams VL, Yu Q, et al. Current patient perspectives of vulvovaginal candidiasis: incidence, symptoms, management and post-treatment outcomes. BMC Women’s Health. 2019;19(1):48.

11. Lema VM. Recurrent vulvo-vaginal candidiasis: diagnostic and management challenges in a developing country context. Obstet Gynecol Int J. 2017;7(5):260.

12. Opoku R, Yar DD, Botchwey CO. Self-medication among pregnant women in Ghana: A systematic review and meta-analysis. Heliyon. 2022;8(10):e10777.

13. Khaskheli M, Baloch S, Baloch AS, Shah SGS. Vaginal discharge during pregnancy and associated adverse maternal and perinatal outcomes. Pakistan journal of medical sciences. 2021;37(5):1302–8.

14. Humaid A, Alghalibi SMS, Al-Mahbashi A, AL-Arossi A, Edrees WH, editors. The Prevalence Of Vulvovaginal Candidiasis In Pregnant Women Attending Several Hospitals In Sana’a, Yemen 2020.

15. Dou N, Li W, Zhao E, Wang C, Xiao Z, Zhou H. Risk factors for candida infection of the genital tract in the tropics. African health sciences. 2014;14(4):835–9.

16. Shukla A, Sobel JD. Vulvovaginitis Caused by Candida Species Following Antibiotic Exposure. Current Infectious Disease Reports. 2019;21(11):44.

17. Eraky EM. The Effect of Supportive Nursing Instructions on Recurrence of Vulvovaginal Candidiasis infection during Pregnancy. 2018.

18. Jombo G, Akpera M, Hemba S, Eyong K. Symptomatic vulvovaginal candidiasis: knowledge, perceptions and treatment modalities among pregnant women of an urban settlement in West Africa. African Journal of Clinical and Experimental Microbiology. 2011;12(1).

